# Prescribing differences among older adults with differing health cover and socioeconomic status: a cohort study

**DOI:** 10.1101/2023.03.30.23287967

**Authors:** Ciaran Prendergast, Michelle Flood, Logan T. Murry, Barbara Clyne, Tom Fahey, Frank Moriarty

## Abstract

**Introduction:** As health reforms move Ireland from a mixed public-private system toward universal healthcare, it is important to understand variations in prescribing practice for patients with differing health cover and socioeconomic status. This study aims to determine how prescribing patterns for patients aged ≥65 years in primary care in Ireland differ between patients with public and private health cover.

**Methods:** This was an observational study using anonymised data collected as part of a larger study from 44 general practices in Ireland (2011-2018). Data were extracted from electronic records relating to demographics and prescribing for patients aged ≥65 years. The cohort was divided between those with public health cover (via the General Medical Services (GMS) scheme) and those without. Standardised rates of prescribing were calculated for pre-specified drug classes. We also analysed the number of medications, polypharmacy, and trends over time between groups, using multilevel linear regression adjusting for age and sex.

**Results:** Overall, 42,456 individuals were included (56% female). Most were covered by the GMS scheme (62%, n=26,490). The rate of prescribing in all medication classes was higher for GMS patients compared to non-GMS patients, with the greatest difference in benzodiazepine anxiolytics. The mean number of unique medications prescribed to GMS patients was 10.9 (SD 5.9), and 8.1 (SD 5.8) for non-GMS patients. The number of unique medications prescribed to both GMS and non-GMS cohorts increased over time. The increase was steeper in the GMS group where the mean number of medications prescribed increased by 0.67 medications/year. The rate of increase was 0.13 (95%CI 0.13, 0.14) medications/year lower for non-GMS patients, a statistically significant difference.

**Conclusion:** Our study found a significantly larger number of medicines were prescribed to patients with public health cover, compared to those without. Increasing medication burden and polypharmacy among older adults may be accelerated for those of lower socioeconomic status. These findings may inform planning for moves towards universal health care, and this would provide an opportunity to evaluate the effect of expanding entitlement on prescribing and medicines use.

## Introduction

With changing population profiles and increasingly costly medical interventions, high and middle-income countries are facing challenges in providing affordable healthcare to their populations.[1] By 2041, citizens over 65 years of age will make up 22% the Irish population; a doubling of the 2006 figure.[2] Delivering functional and affordable systems of universal healthcare requires identifying the optimal healthcare system which balances patients’ needs with services and costs covered.[3] In Ireland, political discussions surrounding healthcare reform culminated in the Sláintecare report in 2017, which provided a roadmap to a future single-payer system of universal healthcare, based on need and not on ability to pay.[4]

At present, the Irish healthcare system is two-tiered and incorporates a mix of both public and private elements.[5] Notably, access to prescription medicines varies considerably for individuals based on income and age. Some patients with full public health cover pay only a small prescription charge for each medicine. Alternatively, individuals who do not meet income and age criteria pay out-of-pocket for the cost of their prescription medicines, up to a monthly household cap. Differences in prescription medicine use between these groups may arise due to differing individual characteristics (i.e. socioeconomic status), but also the effect of differing healthcare.

Existing literature has identified variation in medication prescribing for individuals with public and private health cover and access. Previous studies in countries in Africa and Sweden found physicians working in the private sector are less likely to adhere to guidelines, while also being less likely to prescribe rationally for certain conditions.[6, 7] In Ireland, polypharmacy and potentially inappropriate prescribing have increased in recent years; however, the evidence for prescribing variation between the public and private sector is mixed.[8] A 2008 study found evidence of no difference in prescribing rates, but higher inappropriate prescribing of antipsychotics to individuals in private residential care settings compare to public.[9] A more recent study identified that public patients in Ireland had a 21-38% greater risk of polypharmacy compared to patients with private healthcare coverage. The study authors concluded that publicly funded healthcare in Ireland led to greater medication use in people aged 50-69 years.[10]

International evidence has also examined prescription practices, and in several Swedish studies, private providers were found to prescribe a higher number of medicines, though less cost-effectively, than public GPs.[11, 12] The majority of the studies comparing prescription in the public and private sectors have been carried out with regard to low- or middle-income countries, where a series of comprehensive meta-analyses support the idea that there is measurable variability in prescribing practice between sectors.[13, 14] The Irish system presents a unique opportunity to evaluate prescribing differences among patients with differing healthcare entitlements, cared for by the same providers. An understanding of differences in prescribing patterns between public and private patients in Irish general practice is important if future health reform extends coverage of prescription medicines entitlement.

## Aim and objectives

This study aims to determine how prescribing practices for patients aged 65 years and over in primary care in Ireland differ between patients with public and private health cover.

The objectives are to assess differences in the:

- Rate of prescribing of common drug classes.
- Prevalence of individual drugs within common drug.
- Number of medications prescribed.

## Methods

### Study design, population, and setting

This was an observational study reported in line with the STrengthening the Reporting of OBservational studies in Epidemiology (STROBE) statement.[15] Anonymised data were collected as part of a larger study from 44 general practices in the Republic of Ireland using the patient management software Socrates (www.socrates.ie) between January 2011 and April 2018. Ethical approval was obtained from the Irish College of General Practitioners. Participating practices from the catchment areas of Dublin (n=30), Galway (n=11), and Cork (n=3) hospitals represented 91% of those contacted. Ireland has a mixed public-private health system, and a proportion of the population are entitled to public health cover, with eligibility based on household income and age. The General Medical Service (GMS) scheme covers the most socioeconomically deprived people, approximately one third of the population, and entitles them to GP visits and a range of health services free at the point of access, and prescription medications (with a small co-payment of €2.50).[16] The Doctor Visit Card (DVC) scheme covers people with higher, but still limited, means, who are entitled to free GP visits but pay for other health services and their medications. All other individuals pay for healthcare and prescription medications (with a household cap of €144 per month applying during the study period).

Data were extracted from the patient management system relating to demographics, consultations, prescribing and hospitalisations for patients aged 65 years and older. Patients were included in the present analysis if they had prescriptions issued on at least two dates during the study period, and had demographics (age and sex) and date of prescribing data recorded. Observations with a date of prescription outside of the study period were removed from the analysis.

### Study variables

Prescription records in the dataset are at the medication level and included date of prescription, number of issues (i.e., how many times a prescription could be dispensed), product name, and generic name. Medications were coded using the Anatomical Therapeutic Chemical (ATC) classification, a system developed by the World Health Organisation for drug utilisation research and monitoring. ATC codes are organised by physiological system and are hierarchical, with the full seven-character ATC code identifying the active substance, and the five-character ATC code identifying the chemical subgroup level (usually equivalent to the drug class). Age and sex were extracted as demographic variables from the GP records, as was the type of health cover a patient had: GMS scheme (considered “public”), DVC scheme, or neither of these (considered to be “private”). We grouped DVC scheme cohort with the private cohort as a “non-GMS” category, as although GP visits are covered by the state, medications are not in this instance. We also created a time-varying variable, counting the number of hospitalisations each individual had during the study period.

We calculated the rate of prescribing for drug classes at the five-character ATC code (ATC5) level, both overall and separately for GMS and non-GMS patients. We pre-specified 12 drug classes of interest before commencing the study (Table 1), based on their high prevalence of use, their inclusion in Ireland’s Preferred Drugs Initiative (Health Service Executive Medicines Management Programme),[17] or potential for sub-optimal prescribing.

**Table 1.**
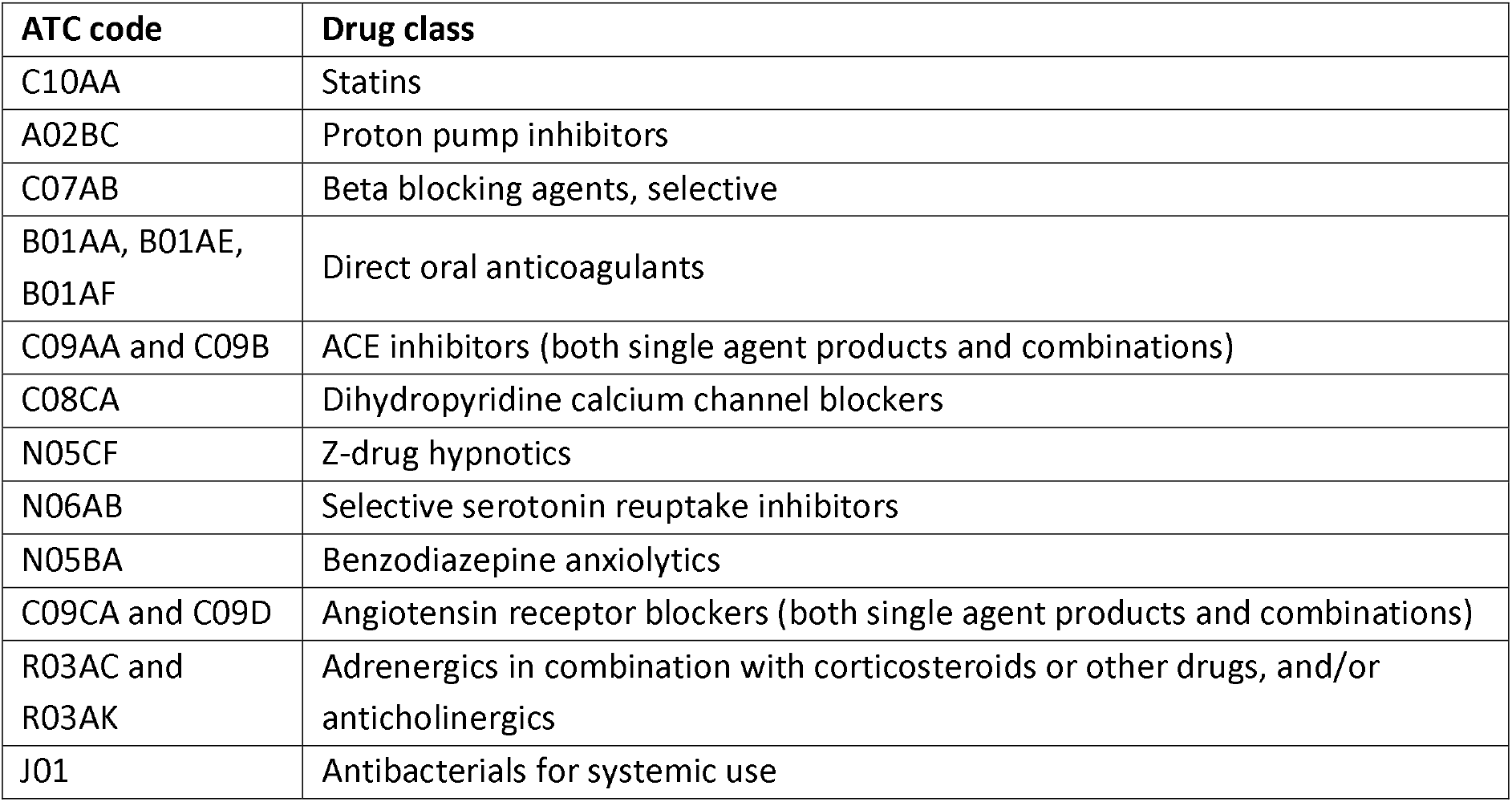
Pre-specified drug classes of interest and corresponding ATC codes.

| ATC code | Drug class |
| --- | --- |
| C10AA | Statins |
| A02BC | Proton pump inhibitors |
| C07AB | Beta blocking agents, selective |
| B01AA, B01AE, B01AF | Direct oral anticoagulants |
| C09AA and C09B | ACE inhibitors (both single agent products and combinations) |
| C08CA | Dihydropyridine calcium channel blockers |
| N05CF | Z-drug hypnotics |
| N06AB | Selective serotonin reuptake inhibitors |
| N05BA | Benzodiazepine anxiolytics |
| C09CA and C09D | Angiotensin receptor blockers (both single agent products and combinations) |
| R03AC and R03AK | Adrenergics in combination with corticosteroids or other drugs, and/or anticholinergics |
| J01 | Antibacterials for systemic use |

We calculated the number of unique drug classes (at the ATC5 level) each patient had been prescribed over the previous 12 months on a rolling basis across the study period, which was used as the number of medicines each patient was prescribed. The number of medicines prescribed was also converted into a categorical variable with prescription of 5-9 medications being classed as ‘polypharmacy’ and 10 or more medications being classed as ‘major polypharmacy’.

### Statistical analysis

First, we described patient characteristics, both overall and separately for GMS and non-GMS patients. We then directly standardised rates of prescribing (based on number of prescriptions and number of repeats/issues per prescription) for drug classes among GMS patients to the non-GMS population, using age group (65-69, 70-74, 75-79, 80-84, 85-89 and 90 years and over) sex, and calendar year, generating 95% confidence intervals (95% CIs) for the rates in both groups. Including year as a standardisation variable accounted for the amount of time patients were present in the dataset. The ratio of the prescribing rate for each drug class among the GMS versus non-GMS patients was plotted as a bubble graph. The same analysis was carried out comparing the GMS group to the DVC group alone, and the private group alone. We determined the prevalence of individual medications (seven-character ATC codes) within each drug class of interest, and assessed any difference between health cover groups in the distribution of prescribing within drug classes using a chi-squared test. A single practice, which was missing number of repeats/issues data, was excluded from this drug class analysis.

We used monthly values for the number of unique drug classes (at the ATC5 level) each patient had been prescribed over the previous 12 months to plot the mean number over time for GMS and non-GMS patients. We also plotted the proportion of GMS and non-GMS patients with polypharmacy over time in categories of 1-4, 5-9, 10-14 and 15+ medications. We also summarised the mean number of medicines prescribed per person over the full study period for the GMS and non-GMS groups, taking an average of the number of medicines each time a prescription was issued (excluding observations in the 12 months after the first date of prescription for an individual, where a full 12-month period for calculating number of medicines was not yet available). We used a multilevel linear regression analyses to assess whether the number of medications differed by health cover and over time. Data was hierarchical with monthly time points, nested within individual patients, nested within GP practices. The fixed covariates included date of prescription (scaled to 1 unit per year and continuous), health cover type (categorical, GMS and non-GMS), age (continuous in years) and sex (categorical, male and female). Random intercepts were included for the patient and practice level, and variance and variance partition coefficients were estimated for each level. A second model was also fitted to include an interaction between date of prescription and health cover, assessing whether any change in number of medicines prescribed over time differed according to health cover. A third model included a hospitalisations variable, to examine how this may explain differences in the number of medications between health cover groups. When modelling, the average number of unique medications prescribed to individuals over time, observations occurring less than 12 months after the first for an individual were removed as incomplete 12-month periods. Analyses were conducted using the lme4 package in R,[18, 19] and statistical significance was assumed at p<0.05.

## Results

The analyses included data on 42,456 individuals, of which 44% (n=18,695) were male and 56% (n=23,761) were female. The majority (62%, n=26,490) of individuals were covered by the GMS scheme, while the remaining 15,966 were non-GMS (70% Private and 30% DVC). The mean age of the GMS cohort was 78.9 years (SD 8.1) and the mean age of the non-GMS cohort was 79.4 (SD 9.2). There was a higher proportion of females in the GMS group (58%) compared to the non-GMS group (52.7%). Demographics and health cover status for participants are included in Table 2.

**Table 2.** Descriptive characteristics of included participants

| Characteristic | Total (n=42,456) | GMS (n=26,490) | Non-GMS (n=15,966) |
| --- | --- | --- | --- |
| Age (years), mean (SD) | 79.0 (8.3) | 78.9 (8.1) | 79.4 (9.2) |
| Age group, n (%) |  |  |  |
| 65-69 years | 7,965 (18.8%) | 3,591 (8.5%) | 4,374 (10.3%) |
| 70-74 years | 9,070 (21.4%) | 5,232 (12.3%) | 3,838 (9.0%) |
| 75-80 years | 7,729 (18.2%) | 5,328 (12.6%) | 2,401 (5.7%) |
| 80-84 years | 6,919 (16.3%) | 5,057 (11.9%) | 1,862 (4.4%) |
| 85-89 years | 5,480 (12.9%) | 3,916 (9.2%) | 1,564 (3.7%) |
| 90+ years | 5,294 (12.5%) | 3,366 (7.9%) | 1,928 (4.5%) |
| Female, n (%) | 23,761 (56.0%) | 15,353 (36.2%) | 8,408 (19.8%) |
| Male, n (%) | 18,695 (44.0%) | 11,137 (26.2%) | 7,558 (17.8%) |
| Health cover, n (%) |  |  |  |
| General Medical Services scheme | 26,490 (62.4%) | 26,490 (100.0%) | 0 |
| Doctor Visit Card | 4,743 (11.2%) | 0 | 4,743 (29.7%) |
| Private | 11,223 (26.4%) | 0 | 11,223 (70.3%) |

### Drug class prescribing

The rate of prescribing in all pre-specified drug classes was higher for GMS patients compared to non-GMS patients. Figure 1 shows the ratios of GMS to non-GMS prescribing rates for these classes. In all cases, the rate of prescribing was at least 1.3 times higher in the GMS group, with the smallest difference in systemic antibacterials. We saw the greatest disparity in benzodiazepine anxiolytics where the rate of GMS prescribing was 1.78 times higher; a rate of 996 per 1000 person-years in the GMS group versus a rate of 559 per 1000 person-years in the non-GMS group. The next largest difference was inhaled adrenergic medication combined with corticosteroids and/or anticholinergics, with a rate 1.58 times higher in the GMS group. Crude and standardised rates for each medication class in each group are reported in supplementary table 1. In sensitivity analysis, ratios of GMS to DVC rates were higher than the corresponding ratio of GMS to private rates in most cases, with the exception of statins, angiotensin receptor blockers, and dihydropyridine calcium channel blockers (Supplementary figure 1). A further sensitivity analysis considering prevalence (i.e. number of people prescribed the medication class, rather than the rate of prescribing per 1,000 person-years) again showed higher prevalence in the GMS group versus non-GMS across all drug classes. The difference were more modest, ranging from 1.04 to 1.30, the largest difference being in inhaled adrenergic combinations (Supplementary figure 2 and supplementary table 2).

**Figure 1:**
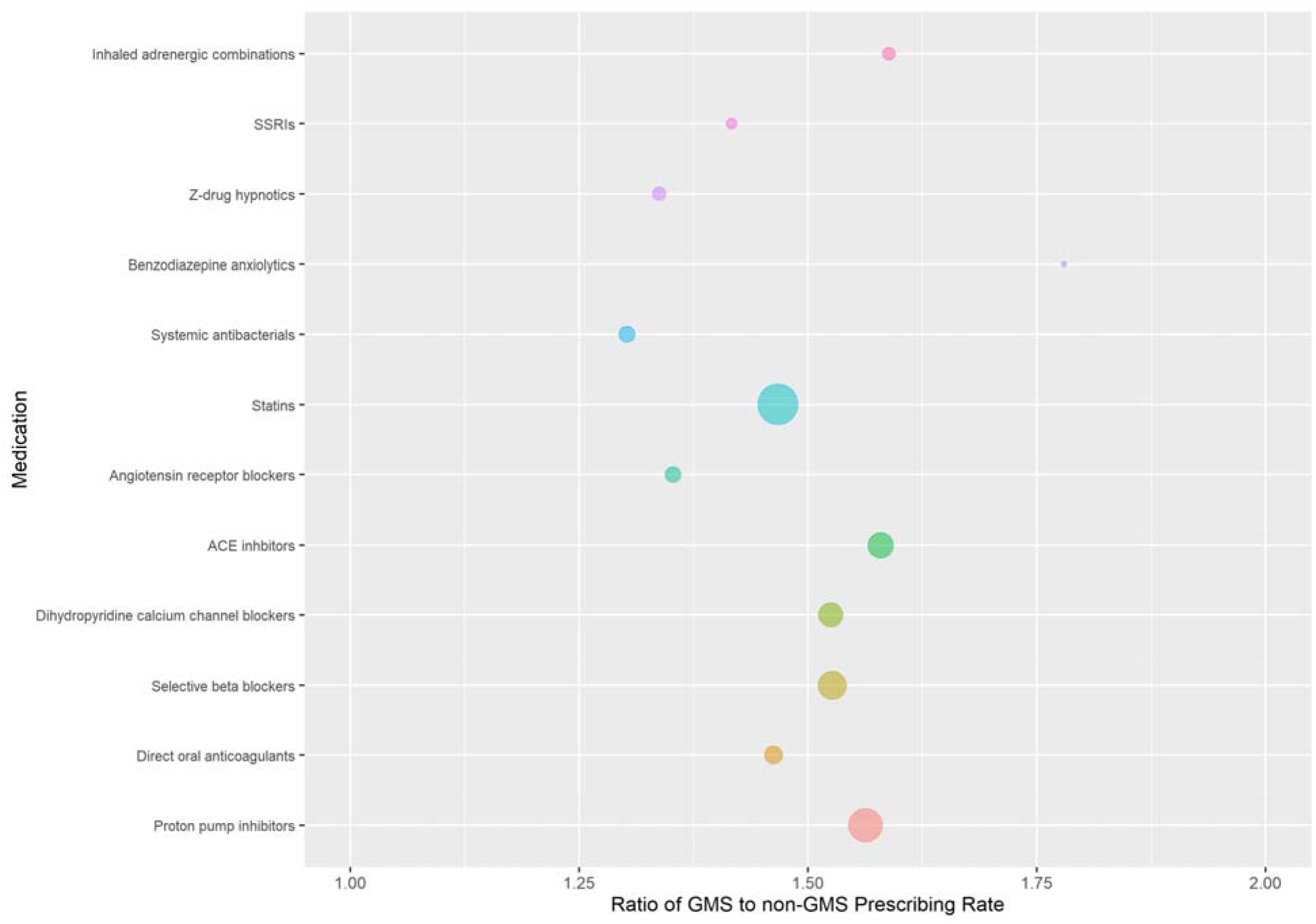
Ratio of GMS to non-GMS prescribing rates for pre-specified medication classes, with bubble size indicating the rate of prescribing of each class among GMS patients

As examples, the mosaic plots below (figure 2) show the relative proportions of medications (at the ATC7 level) that make up four of the pre-specified drug classes (benzodiazepine anxiolytics, statins, inhaled adrenergic combinations, and calcium channel blockers). For benzodiazepine anxiolytics, diazepam makes up a significantly greater proportion of prescribing in the GMS group compared to the non-GMS group (a difference of 5 percentage points), whereas the reverse is true of alprazolam (which is 2 percentage points higher among non-GMS patients). Within calcium channel blockers, amlodipine makes up a significantly greater proportion of prescribing within the non-GMS cohort (a difference of 4 percentage points). For inhaled adrenergic combinations, salmeterol/fluticasone made up significantly more prescribing in the GMS group (4.5 percentage points higher), whereas formoterol/budesonide made up significantly more of non-GMS group prescribing for this drug class (5 percentage points higher). For statins, the largest difference was rosuvastatin accounting for 3 percentage points more of statin prescribing in the non-GMS group. Mosaic plots for the other drug classes are included as Supplementary figure 3, and frequency tables for medications within each drug class by health cover are included as supplementary table 3.

**Figure 2:**
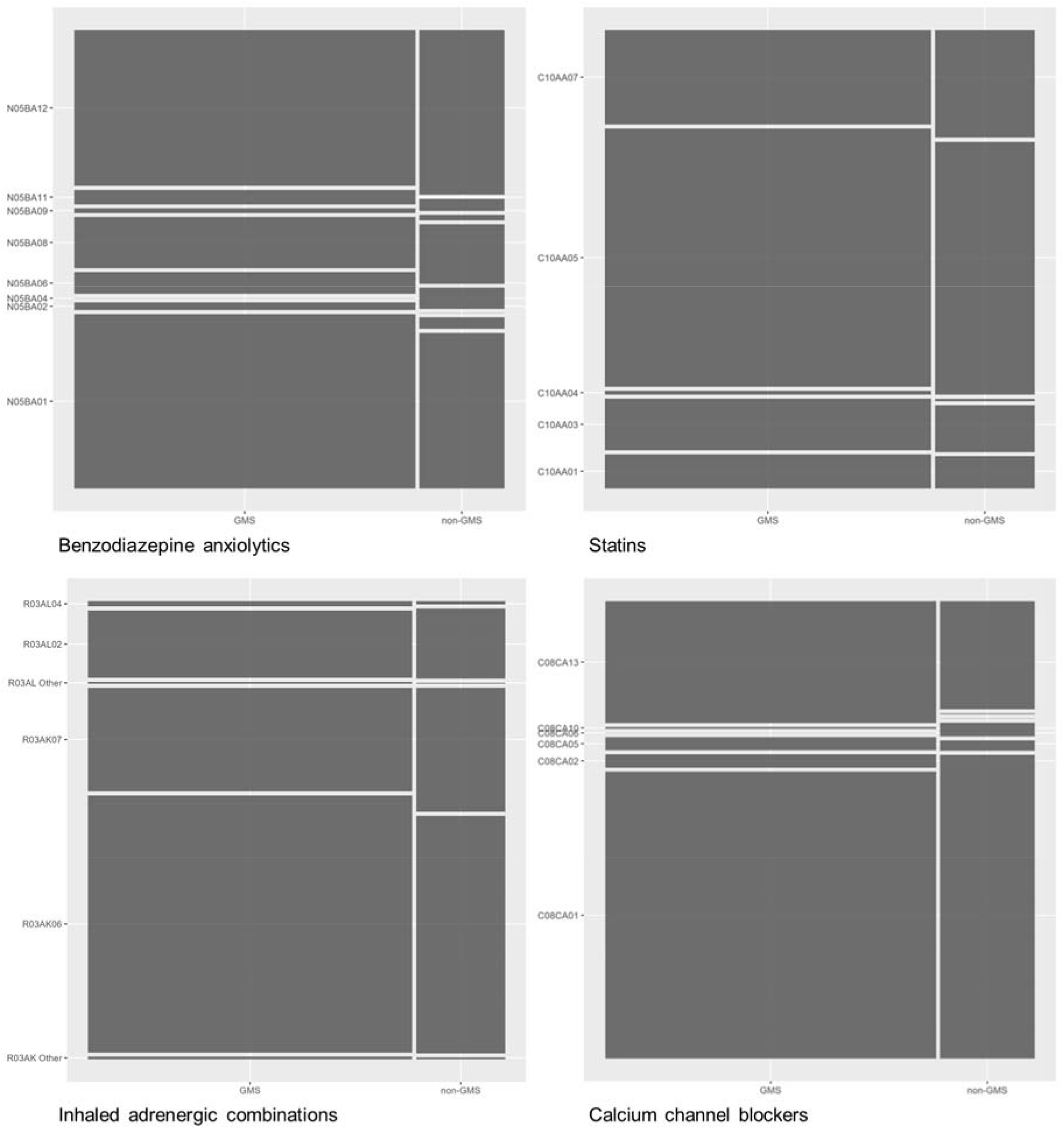
The relative proportions of individual medication prescribing (indicated by ATC7 codes) for (clockwise from top left) benzodiazepine anxiolytics, statins, dihydropyridine calcium channel blockers, and inhaled adrenergic combinations in the GMS and non-GMS groups

### Number of medications

The number of unique medications prescribed to both the GMS and non-GMS cohorts increased over time, as depicted by the time trend below. The increase was more pronounced and more sustained in the GMS group, rising from a mean of 7.3 (SD 5.8) medications in January 2011 to a level of 14.2 (SD 7.1) in April 2018 compared to the non-GMS group rising from 5.8 (SD 4.8) to 9.2 (SD 6.6). Figure 3, shows the fitted line for the number of medicines over time for each group.

**Figure 3:**
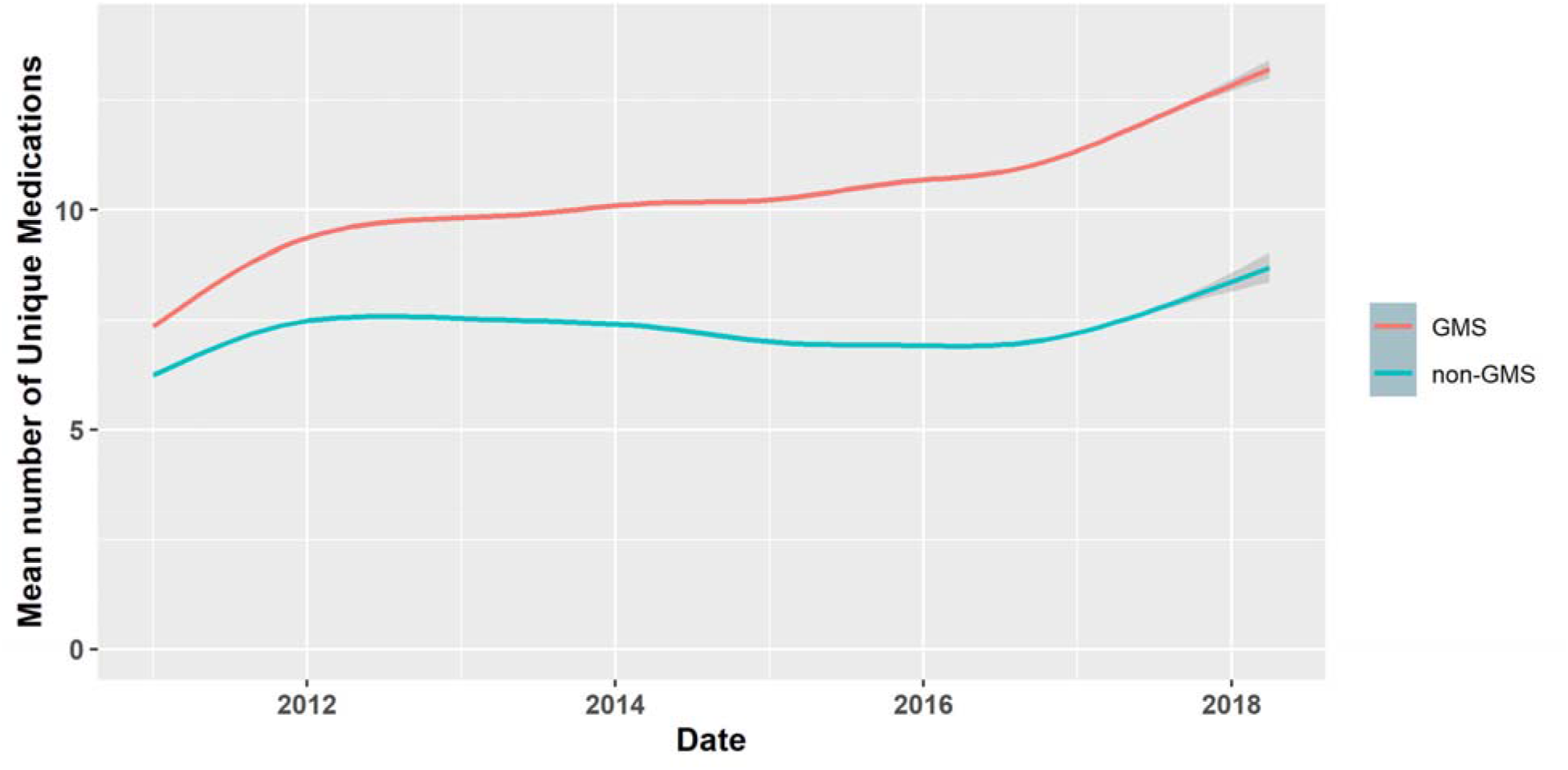
Time trend comparing the changes in number of unique medications prescribed to both GMS and non-GMS groups over time, with grey shading indicated 95% confidence intervals.

The rates of polypharmacy (≥5 medications), and major polypharmacy (≥10 medications) over time are shown in the stacked area chart below. The GMS group began the study period with higher rates of major polypharmacy and this became more pronounced over time. The rate of major polypharmacy in the GMS group increased from 33.2% in January 2011 to 76.5% in April 2018.

The mean number of unique drug classes prescribed to GMS patients over the full study period was 10.9 (SD 5.9), compared to a mean of 8.1 (SD 5.8) among non-GMS patients. Similarly, the median number of unique drug classes prescribed (Figure 5) was higher among GMS patients at 10.1 (IQR 6.5 to 14.3) compared to non-GMS patients (median 6.6, IQR 3.7 to 11.1).

**Figure 4:**
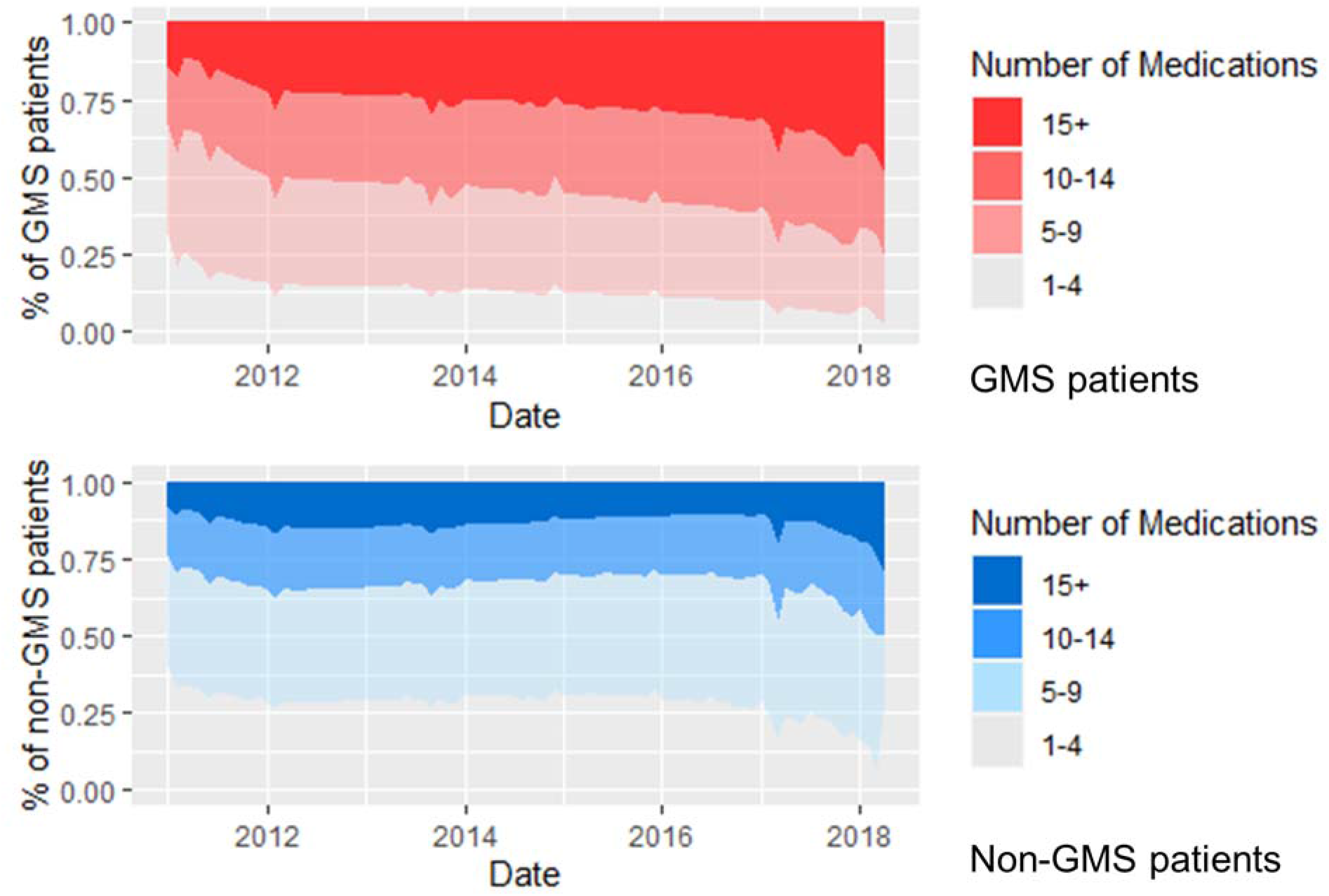
Proportion of patients with levels of polypharmacy over the study period for GMS (top) and non-GMS groups (bottom)

**Figure 5:**
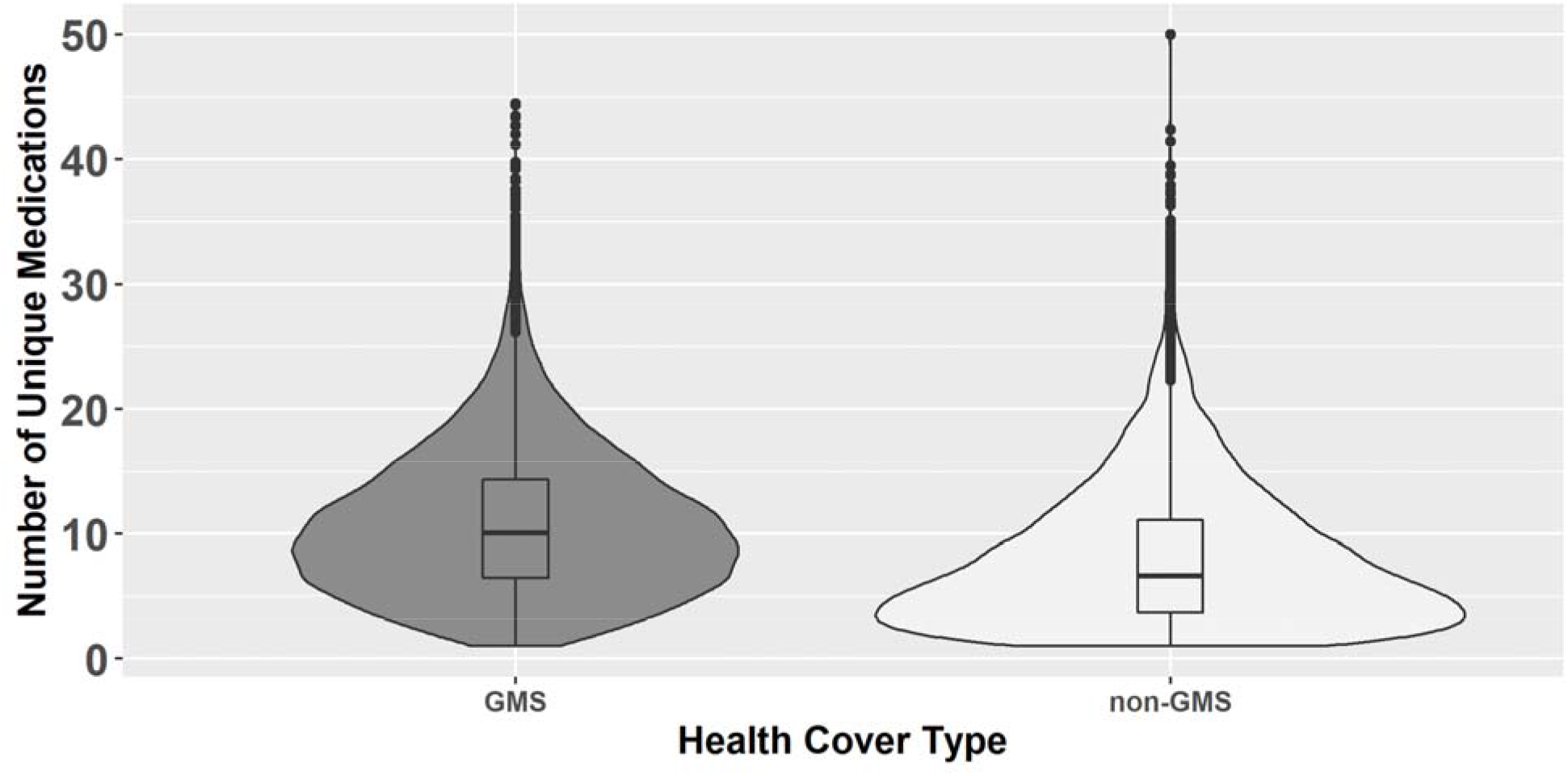
Violin plot showing the number of unique medications prescribed to GMS and non-GMS patients.

The results of the multilevel regression model are shown in Table 3. Based on variance partition coefficients in Model 1, 4% of variation was between practices, 75% was between patients within practices, and 21% was within patients over time. There was a statistically significant increase in number of unique medications over time (0.65 additional medicines per year, 95% CI 0.64, 0.65), with non-GMS patients (compared to GMS patients) being prescribed 1.93 (95% CI 2.00, 1.87) fewer medications. Being female was associated with a higher number of medicines (0.91 additional medicines, 95% CI 0.85, 0.96) compared to males. In model 2, including an interaction term between time and health cover, the VPC were similar to model 1. In this model, mean number of medications prescribed increased by 0.67 medications/year for GMS patients. The rate of increase was 0.13 (95%CI 0.13, 0.14) medications/year lower for non-GMS patients, a statistically significant difference. In model 3, including a variable counting the number of hospitalisations, the increase in medications over time and the difference in the rate of increase by health cover were both attenuated.

**Table 3:**
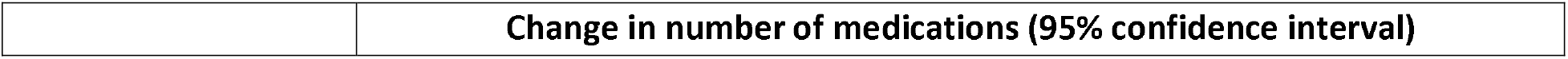
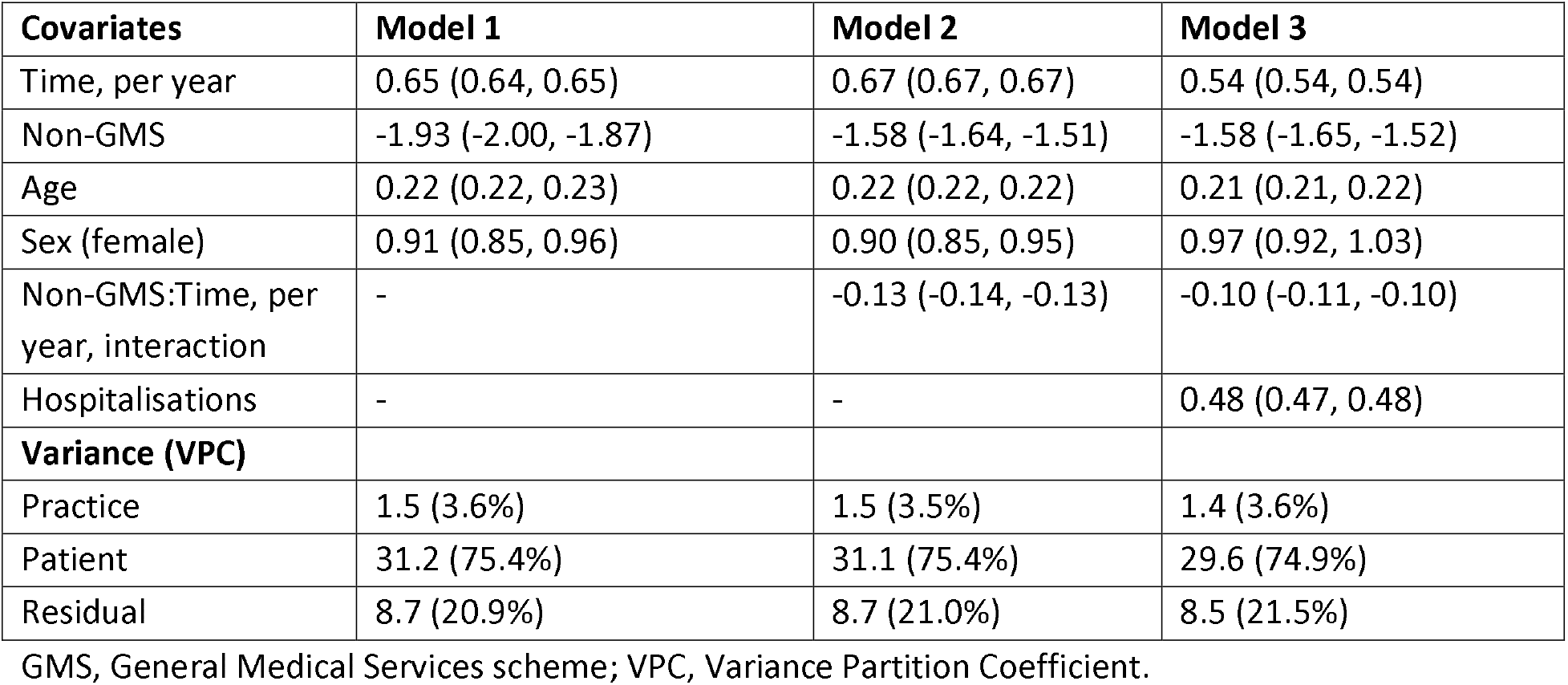
Characteristics associated with number of unique medications over time in multilevel linear regression

|  | Change in number of medications (95% confidence interval) |
| --- | --- |

| **Covariates** | **Model 1** | **Model 2** | **Model 3** |
| --- | --- | --- | --- |
| Time, per year | 0.65 (0.64, 0.65) | 0.67 (0.67, 0.67) | 0.54 (0.54, 0.54) |
| Non-GMS | -1.93 (-2.00, -1.87) | -1.58 (-1.64, -1.51) | -1.58 (-1.65, -1.52) |
| Age | 0.22 (0.22, 0.23) | 0.22 (0.22, 0.22) | 0.21 (0.21, 0.22) |
| Sex (female) | 0.91 (0.85, 0.96) | 0.90 (0.85, 0.95) | 0.97 (0.92, 1.03) |
| Non-GMS:Time, per year, interaction | - | -0.13 (-0.14, -0.13) | -0.10 (-0.11, -0.10) |
| Hospitalisations | - | - | 0.48 (0.47, 0.48) |
| **Variance (VPC)** |  |  |  |
| Practice | 1.5 (3.6%) | 1.5 (3.5%) | 1.4 (3.6%) |
| Patient | 31.2 (75.4%) | 31.1 (75.4%) | 29.6 (74.9%) |
| Residual | 8.7 (20.9%) | 8.7 (21.0%) | 8.5 (21.5%) |

GMS, General Medical Services scheme; VPC, Variance Partition Coefficient.

## Discussion

In this study, we found higher numbers of medications prescribed to older adults with public health cover (GMS) compared to those without. We also identified steeper growth in the number of medications over time within the GMS cohort. This is reflected in higher rates of prescribing of all of the pre-specified medication classes we examined, with the greatest difference in rates for inhaled adrenergic combination medications. Within drug classes, there were some differences in the percentage share of individual medications between health cover groups, however these did not consistently align with national preferred drug guidance. The steeper growth in medications over time for the GMS group was partly explained by the higher rate of hospitalisation.

Direct comparison with other research is challenging, as most examine prescribing differences between patients attending public versus private providers in other healthcare systems, rather than the same providers prescribing to those with differing healthcare entitlements, as in Ireland’s health system. Studies by Granlund (2009) and Hakansson et al. (2001) found a significantly larger number of unique medicines were prescribed to public, rather than private patients. In the Irish setting, Mohan et al. (2021) also reported this disparity in number of medications and the faster growth over time in the over 50s public cohort in Ireland.[11, 12, 20]

One reason for the disparity we identified may be that socioeconomic status is known to correlate negatively with several measures of health, in Ireland.[21] As a result, the publicly covered population is likely to have a higher illness burden, requiring greater pharmaceutical intervention. This association is robust in the literature as made clear by Pathirana and Jackson, (2018), who performed a systemic review encompassing of 24 cross-sectional studies, primarily in high-income settings, showing level of educational attainment and deprivation (as measures of socioeconomic status) were both associated with increased risk of multimorbidity.[22] Guthrie et al. (2015) identified an association between living in a deprived area and increasing polypharmacy among adults of all ages in a region of Scotland.[23] Also in Scotland, a study by Barnett et al. (2012) showed that the accumulation of chronic conditions was more substantial, and occurred earlier (by up to 15 years), in those of a lower socioeconomic status.[24] Given the overrepresentation of socioeconomic deprivation among those with public health cover in Ireland, this may partly explain higher rate of growth in medication burden among GMS patients.

Inhaled adrenergic combination medications showed the second largest or largest difference in prescribing (across prescribing rates or prevalence) of our chosen drug classes, which is striking, as there is a particularly strong negative correlation between socioeconomic status and respiratory diseases.[25] Previous evidence in Ireland has shown this relationship, and respiratory diseases as a whole are more common in Ireland than in many comparable developed nations in Europe (O’Shea, 1997).[26] By way of partial explanation, rates of smoking in Ireland have historically been shown to be significantly higher in those of lower socioeconomic status (Layte and Whelan, 2009).[27] The smallest difference in prescribing rates was for systemic antibacterials, being 1.3-fold higher in GMS patients. Unlike most of the other drug classes examined, these are often short-term prescriptions (thus the impact of deprivation on illness burden may be amplified/propagated less). Further evidence from Scotland found an association between deprivation and rates of antimicrobial prescribing.[28] In contrast, a previous study in Ireland including individuals of all ages found private patients were more likely to receive an antibiotic prescription than GMS patients, however this was reversed among patients aged 65 years and over, consistent with our findings.[29] The less pronounced difference in prescribing rates may also be partly explained by the existence of primary care antimicrobial prescribing guidelines in Ireland since 2012.[30]

An increase in medication burden post hospitalisation is a common occurrence.[31, 32] However, whether the increased medication burden is maintained after discharge is often not examined.[33, 34] We addressed this issue with a multilevel regression model that accounted for the association of hospitalisation with number of medications over time, and found a sustained positive effect. The appropriateness of the increased medication burden is unclear. Viktil et al., (2012), cite a similar number of medication changes upon discharge, commenting on a delay in receipt of discharge notes and speculate that failure to communicate between primary and secondary care contributes to potentially inappropriate prescribing. This finding is built upon by Coll et al., (2021), who show that the inclusion of instructions upon discharge accelerates the discontinuation of benzodiazepines and Z-drugs in older adults.[35] Further work by Perez et al., (2018), suggest that the risk of potentially inappropriate prescribing increases with rates of hospitalisation and degree of multimorbidity.[36] Patients were found to be 72% more likely to have been prescribed a potentially inappropriate medication after a single hospitalisation.

However, Corsonello et al. (2007) suggest that due to their finding that the new drugs tended to relate to chronic conditions, they may largely represent a ‘true and stable’ increase. This may be reflected in our study, as the cohort that accrues chronic conditions earlier and to a greater degree, show the largest increase in polypharmacy. Our study did not account for changes to medication regimens that produced no overall change in medication burden, though this has been put forward as an indicator for identifying patients at risk of potentially inappropriate prescribing.[37]

Our study provides a longitudinal analysis of polypharmacy, a view which is under reported in the literature. Falster et al., highlight that although the medications that make up patients’ polypharmacy change regularly over time, once reached, chronic polypharmacy is often permanent among older patients.[38] A limitation of our study is that we were unable to examine which factors relating to public healthcare entitlement (i.e. increased access to healthcare and medications, or the underlying differences in socioeconomic status) have the greatest relationship with prescribing differences. Therefore, it is not possible to conclude what prescribing rates would be if healthcare entitlement was widened. Although we found higher prescribing rates across our pre-specified drug classes, other classes could potentially show different patterns. However our overall findings for number of medications and polypharmacy support a widespread relationship. Our analysis was also limited to those aged 65 years and over, and therefore cannot be generalised to younger patients. However, the older age group account for the majority of medication utilisation.

## Conclusion

Our study found a significantly larger number of unique medicines were prescribed to patients with public health cover, compared to those without. This disparity increased over time and was consistent within all drug classes analysed. This may be driven by socioeconomic deprivation rather than health cover. We provide new evidence that the growth in medication burden and polypharmacy among older adults is accelerated for those of lower socioeconomic status, and evidence to support decisions about extending medications entitlement further in Ireland in the future. Such an expansion would provide a further opportunity to assess the impact of extended entitlement on prescribing and medicines use.

## Declarations

### Ethics approval and consent to participate

Approval was granted by the Irish College of General Practitioners Research Ethics Committee. This is a secondary analysis of data that was anonymised before automatic extraction from electronic records, and therefore individual consent was not required.

### Availability of data and materials

The dataset analysed during the current study is not publicly available as provisions for data sharing were not included in the initial ethical approval.

### Authors’ contributions

MF, BC, TF, and FM conceived the study. CP, MF, TF and FM designed the study. TF provided the data. CP and FM cleaned and analysed the data. All authors interpreted the data. CP and FM drafted the manuscript, and MF, LTM, BC and TF critically revised the manuscript.

## Acknowledgements

We wish to acknowledge the general practitioners and patients whose data was contributed to this study.

## Additional materials

**Supplementary figure 1.**
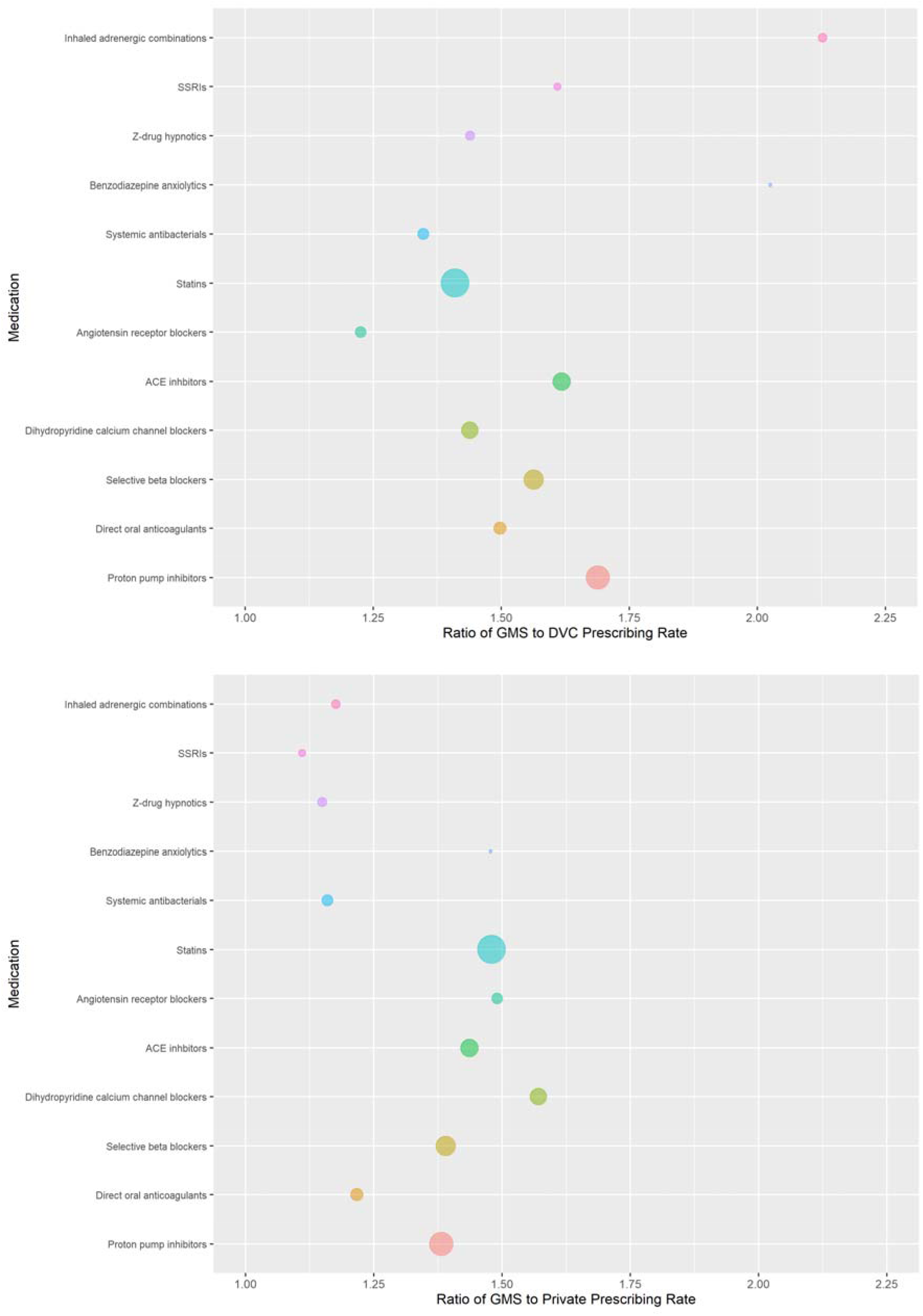
Ratio of GMS to DVC (top) and GMS to private (bottom) prescribing rates for pre-specified medication classes, with bubble size indicating the rate of prescribing of each class among GMS patients

**Supplementary figure 2.**
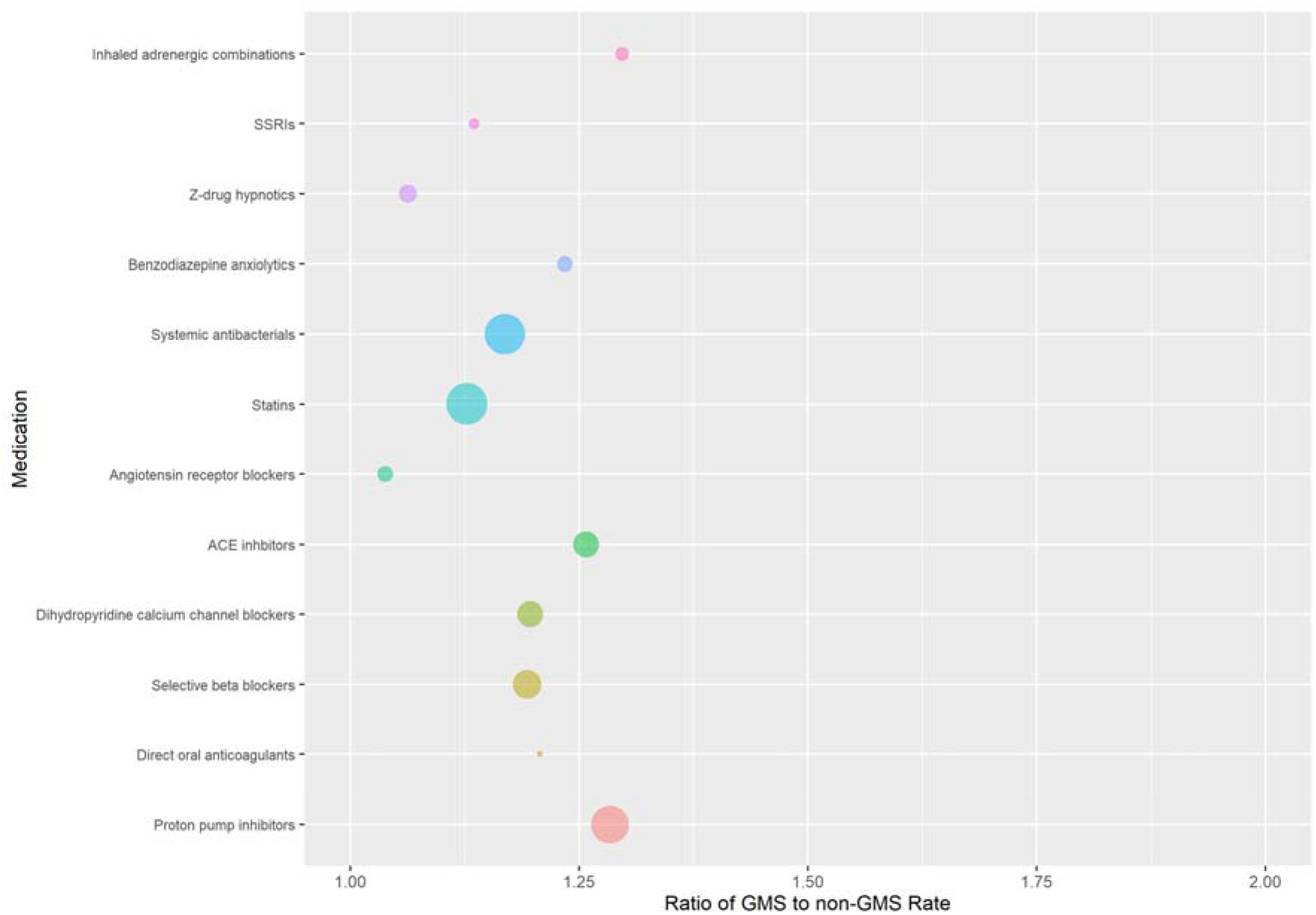
Ratio of GMS to non-GMS prevalence of prescribing (i.e. percentage of individuals with at least one prescription) for pre-specified medication classes, with bubble size indicating the prevalence of each class among GMS patients

**Supplementary figure 3.**
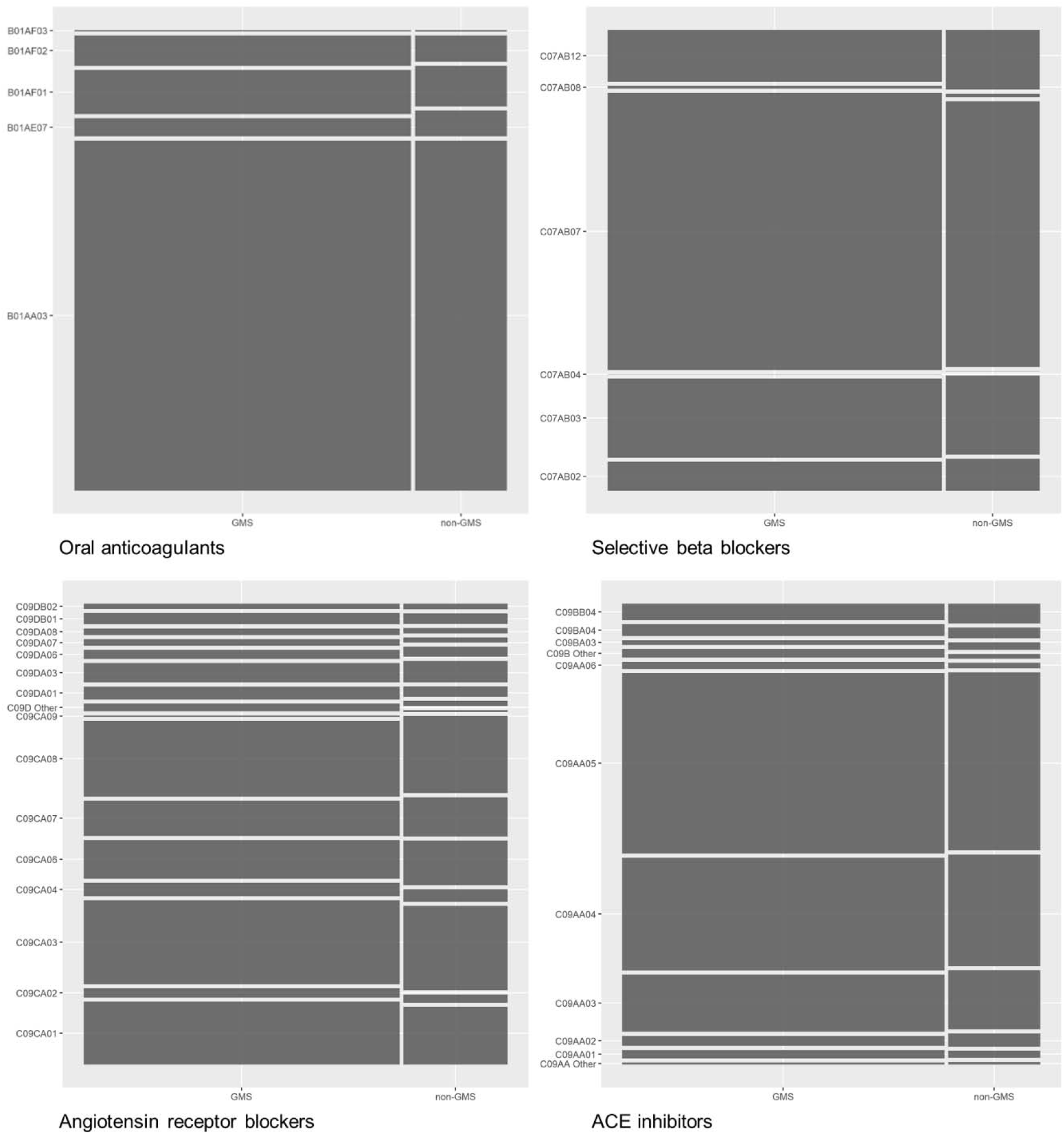
Relative proportions of individual medication prescribing (indicated by ATC7 codes) for cardiovascular drug classes in GMS and non-GMS groups (clockwise from top left oral anticoagulants, selective beta blockers, ACE inhibitors, and angiotensin receptor blockers)

**Supplementary figure 4.**
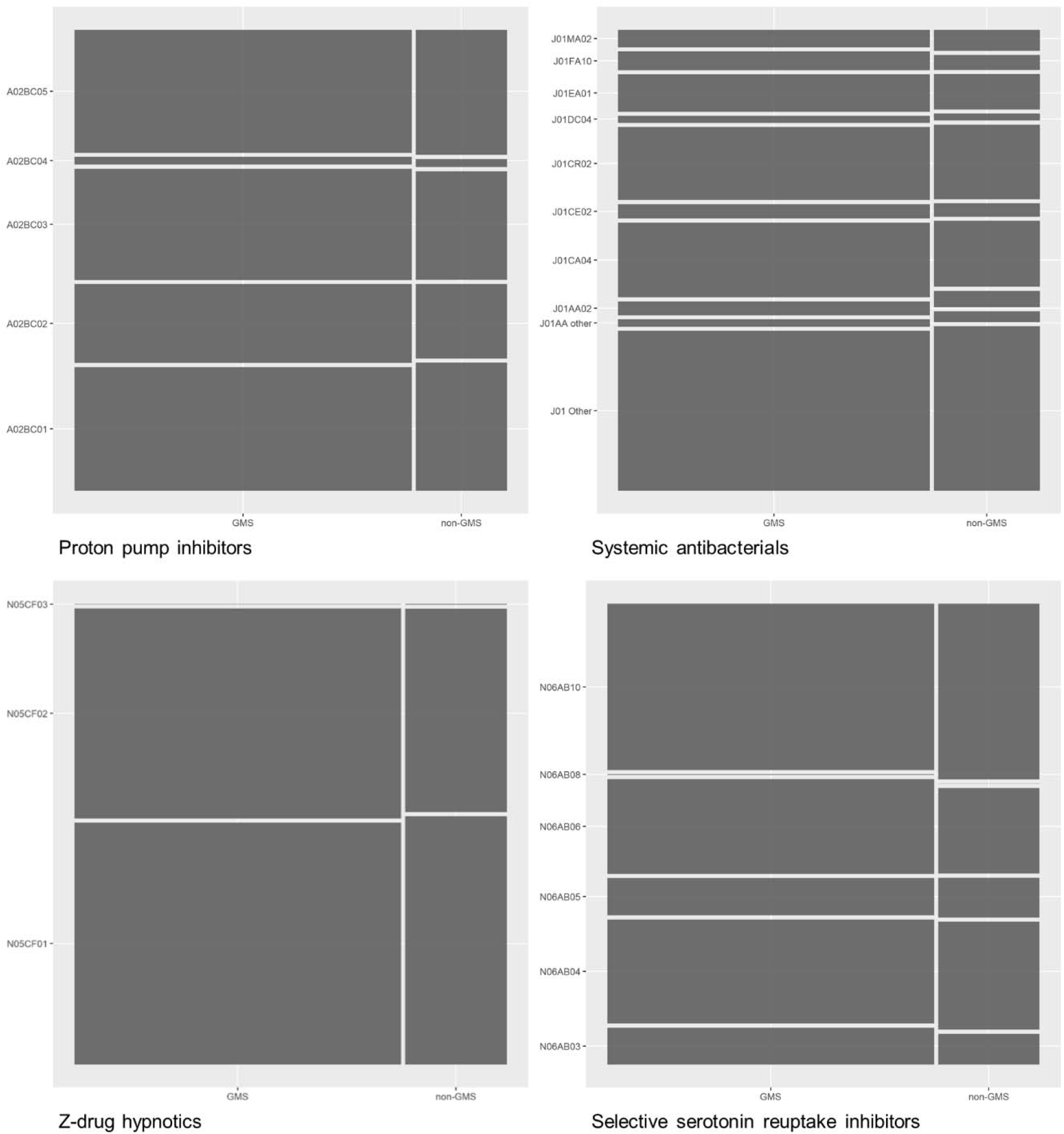
Relative proportions of individual medication prescribing (indicated by ATC7 codes) for other drug classes in GMS and non-GMS groups (clockwise from top left proton pump inhibitors, systemic antibacterials, selective serotonin reuptake inhibitors and Z-drug hypnotics

**Supplementary table 1.** Standardised prescribing rates for pre-specified drug classes in GMS and non-GMS groups anxiolytics

| **Subgroup** | **GMS** |  | **Non-GMS** |  |
| --- | --- | --- | --- | --- |
|  | **Prescriptions** | **Standard rate of prescriptions per 1,000 person years** | **Prescriptions** | **Standard rate of prescriptions per 1,000 person years** |
| PPIs | 586,552 | 4,110 (4,099, 4,120) | 158,142 | 2,630 (2,615, 2,644) |
| ARBs | 204,019 | 1,429 (1,423, 1,436) | 67,089 | 1,057 (1,048, 1,066) |
| SSRIs | 159,771 | 1,119 (1,114, 1,125) | 49,327 | 790 (782, 798) |
| Selective beta blockers | 423,889 | 2,970 (2,961, 2,979) | 118,735 | 1,946 (1,934, 1,958) |
| Systemic antibacterials | 212,795 | 1,491 (1,485, 1,497) | 72,154 | 1,145 (1,136, 1,154) |
| Statins | 787,234 | 5,516 (5,504, 5,528) | 239,687 | 3,759 (3,742, 3,776) |
| ACE inhibitors | 363,001 | 2,543 (2,535, 2,552) | 103,557 | 1,611 (1,600, 1,621) |
| Z-drugs | 181,434 | 1,271 (1,265, 1,277) | 56,383 | 950 (942, 959) |
| Direct oral anticoagulants | 232,610 | 1,630 (1,623, 1,636) | 63,312 | 1,114 (1,105, 1,124) |
| Inhaled adrenergic combinations | 176,463 | 1,236 (1,231, 1,242) | 48,405 | 778 (771, 786) |
| Dihydropyridine CCBs | 341,339 | 2,392 (2,384, 2,400) | 97,381 | 1,568 (1,557, 1,579) |
| Benzodiazepine anxiolytics | 142,097 | 996 (990, 1,001) | 35,295 | 559 (553, 566) |

ACE, angiotensin converting enzyme; ARB, angiotensin receptor blocker; CCB, calcium channel blockers; GMS, General Medical Services; PPI, proton pump inhibitor; SSRI, selective serotonin reuptake inhibitor

**Supplementary table 2.** Standardised prescribing rates for pre-specified drug classes in GMS, DVC and private groups

| Subgroup | GMS |  | DVC |  | Private |  |
| --- | --- | --- | --- | --- | --- | --- |
|  | Prescriptions | Standard rate of prescriptions per 1,000 person years | Prescriptions | Standard rate of prescriptions per 1,000 person years | Prescriptions | Standard rate of prescriptions per 1,000 person years |
| DVC |  |  |  |  |  |  |
| PPIs | 586,552 | 3,801 (3,791, 3,811) | 58,495 | 2,251 (2,230, 2,272) | 99,647 | 2,751 (2,727, 2,775) |
| ARBs | 204,019 | 1,322 (1,316, 1,328) | 30,189 | 1,079 (1,065, 1,093) | 36,900 | 887 (874, 899) |
| SSRIs | 159,771 | 1,035 (1,030, 1,040) | 15,179 | 643 (631, 655) | 34,148 | 933 (919, 947) |
| Selective beta blockers | 423,889 | 2,747 (2,739, 2,755) | 47,608 | 1,757 (1,739, 1,775) | 71,127 | 1,976 (1,956, 1,997) |
| Systemic antibacterials | 212,795 | 1,379 (1,373, 1,385) | 26,046 | 1,023 (1,008, 1,038) | 46,108 | 1,189 (1,173, 1,205) |
| Statins | 787,234 | 5,101 (5,090, 5,113) | 103,155 | 3,618 (3,592, 3,644) | 136,532 | 3,447 (3,421, 3,473) |
| ACE inhibitors | 363,001 | 2,352 (2,345, 2,360) | 40,062 | 1,454 (1,437, 1,471) | 63,495 | 1,638 (1,620, 1,656) |
| Z-drugs | 181,434 | 1,176 (1,170, 1,181) | 20,107 | 817 (804, 830) | 36,276 | 1,023 (1,008, 1,037) |
| Direct oral anticoagulants | 232,610 | 1,507 (1,501, 1,513) | 25,445 | 1,007 (992, 1,021) | 37,867 | 1,238 (1,222, 1,255) |
| Inhaled adrenergic combinations | 176,463 | 1,143 (1,138, 1,149) | 13,943 | 537 (527, 548) | 34,462 | 972 (958, 986) |
| Dihydropyridine CCBs | 341,339 | 2,212 (2,204, 2,219) | 42,557 | 1,537 (1,520, 1,554) | 54,824 | 1,408 (1,392, 1,424) |
| Benzodiazepine anxiolytics | 142,097 | 921 (916, 926) | 11,672 | 455 (445, 464) | 23,623 | 623 (612, 634) |

**Supplementary table 3.**
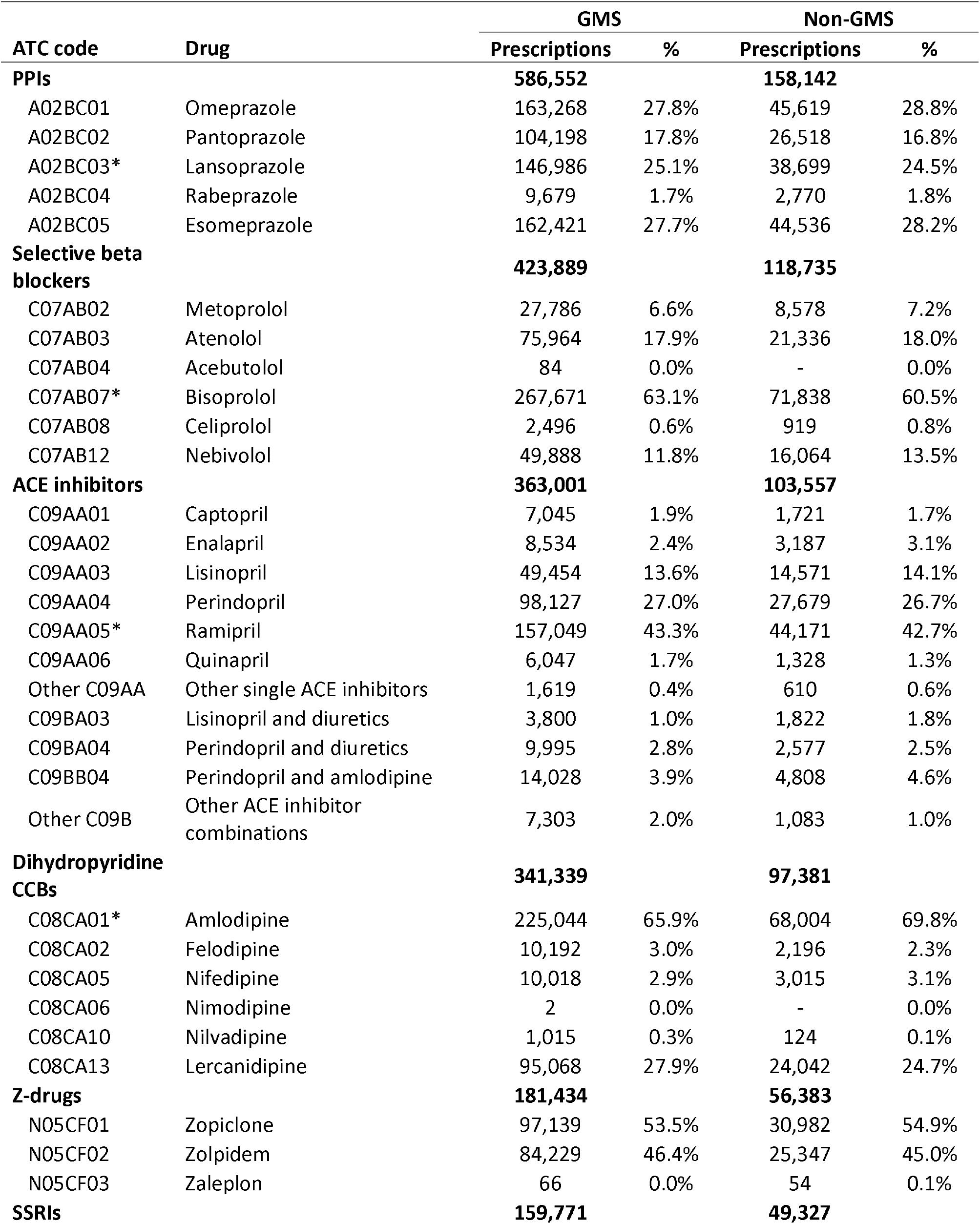
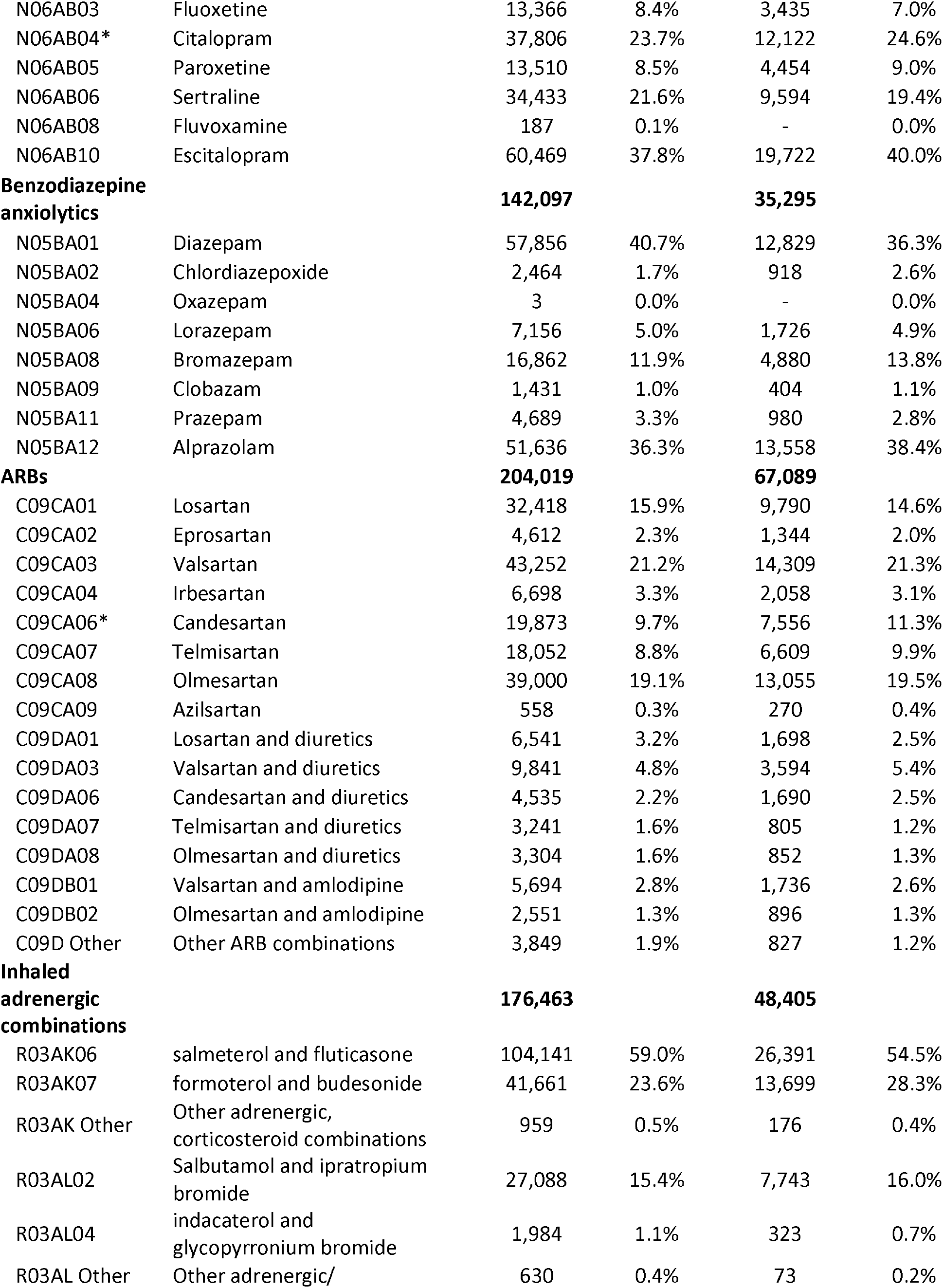
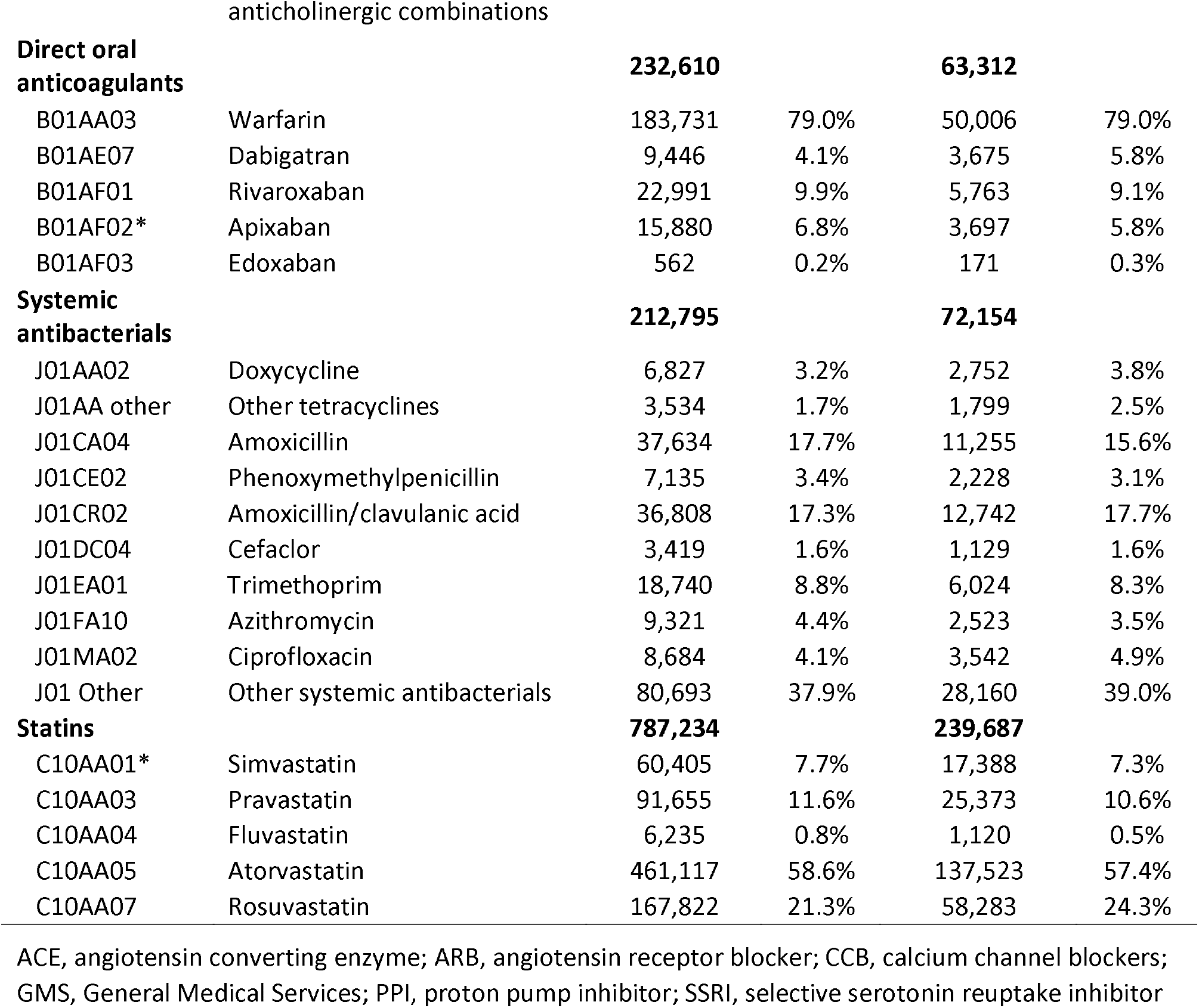
Number and relative proportions of individual medication prescribing (indicated by ATC7 codes) within pre-specified drug classes for GMS and non-GMS groups

## References

1. Vos T, Barber RM, Bell B, Bertozzi-Villa A, Biryukov S, Bolliger I, et al. Global, regional, and national incidence, prevalence, and years lived with disability for 301 acute and chronic diseases and injuries in 188 countries, 1990-2013: A systematic analysis for the Global Burden of Disease Study 2013. Lancet. 2015. https://doi.org/10.1016/S0140-6736(15)60692-4.

2. CSO. Population and Labour Force Projections: 2011–2041. 2008.

3. WHO. Making fair choices on the path to universal health coverage: Final report of the WHO Consultative Group on Equity and Universal Health Coverage. Health Econ Policy Law. 2014.

4. Houses of the Oireachtas Committee on the future of healthcare. Committee on the Future of Healthcare Sláintecare Report. Houses of the Oireachtas, Dublin; 2017.

5. European Observatory on Health Systems and Policies. Ireland: Country Health Profile 2021.

6. Ofori-Asenso R, Brhlikova P, Pollock AM. Prescribing indicators at primary health care centers within the WHO African region: A systematic analysis (1995-2015). BMC Public Health. 2016;16:1–14.

7. Ohlsson H, Merlo J. Is physician adherence to prescription guidelines a general trait of health care practices or dependent on drug type?-A multilevel logistic regression analysis in South Sweden. Pharmacoepidemiol Drug Saf. 2009;18:682–90.

8. Moriarty F, Hardy C, Bennett K, Smith SM, Fahey T. Trends and interaction of polypharmacy and potentially inappropriate prescribing in primary care over 15 years in Ireland: a repeated cross-sectional study. BMJ Open. 2015;5:e008656.

9. Murphy J, O’Keeffe ST. Frequency and appropriateness of antipsychotic medication use in older people in long-term care. Ir J Med Sci. 2008;177:35–7.

10. Richardson K, Kenny RA, Bennett K. The effect of free health care on polypharmacy: A comparison of propensity score methods and multivariable regression to account for confounding. Pharmacoepidemiol Drug Saf. 2014;23:656–65.

11. Hakansson A, Andersson H, Cars H, Melander A. Prescribing, prescription costs and adherence to formulary committee recommendations: Long-term differences between physicians in public and private care. Eur J Clin Pharmacol. 2001;57:65–70.

12. Granlund D. Are private physicians more likely to veto generic substitution of prescribed pharmaceuticals? Soc Sci Med. 2009;69:1643–50.

13. Ofori-Asenso R, Agyeman AA. A review of injection and antibiotic use at primary health care (public and private) centers in Africa. Journal of Pharmacy and Bioallied Sciences. 2015;7:175–80.

14. Basu S, Andrews J, Kishore S, Panjabi R, Stuckler D. Comparative Performance of Private and Public Healthcare Systems in Low- and Middle-Income Countries: A Systematic Review. PLoS Med. 2012;9:e1001244.

15. von Elm E, Altman DG, Egger M, Pocock SJ, Gøtzsche PC, Vandenbroucke JP. The Strengthening the Reporting of Observational Studies in Epidemiology (STROBE) statement: guidelines for reporting observational studies. PLoS Med. 2007;4:e296.

16. Mattsson M, Flood M, Wallace E, Boland F, Moriarty F. Eligibility rates and representativeness of the General Medical Services scheme population in Ireland 2017-2021: A methodological report. HRB Open Res 2022 567. 2022;5:67.

17. McDowell R, Bennett K, Moriarty F, Clarke S, Barry M, Fahey T. An evaluation of prescribing trends and patterns of claims within the Preferred Drugs Initiative in Ireland (2011 – 2016): an interrupted time-series study. BMJ Open. 2018;8:e019315.

18. Bates D, Mächler M, Bolker B WS. “Fitting Linear Mixed-Effects Models Using lme4.” J Stat Softw. 2015;67:1–48.

19. R Core Team. R: A language and environment for statistical computing. R Foundation for Statistical Computing, Vienna, Austria. https://www.R-project.org/. 2021.

20. Mohan G, Nolan A, Moriarty F. The Introduction of Cost Sharing for Prescription drugs: Evidence from The Irish Longitudinal Study of Ageing (TILDA). Econ Soc Rev (Irel). 2021;52:1–40.

21. Teljeur C, Smith SM, Paul G, Kelly A, O’Dowd T. Multimorbidity in a cohort of patients with type 2 diabetes. https://doi.org/103109/138147882012714768. 2013;19:17–22.

22. Pathirana TI, Jackson CA. Socioeconomic status and multimorbidity: a systematic review and meta-analysis. Aust N Z J Public Health. 2018;42:186–94.

23. Guthrie B, Makubate B, Hernandez-Santiago V, Dreischulte T. The rising tide of polypharmacy and drug-drug interactions: population database analysis 1995–2010. BMC Med. 2015;13.

24. Guthrie B, Barnett K, Mercer SW, Norbury M, Watt G, Wyke S. Epidemiology of multimorbidity and implications for health care, research, and medical education: a cross-sectional study. Lancet. 2012;380:37–43.

25. Sahni S, Talwar A, Khanijo S, Talwar A. Socioeconomic status and its relationship to chronic respiratory disease. Adv Respir Med. 2017;85:97–108.

26. O’Shea E. Male mortality differentials by socio-economic group in Ireland. Soc Sci Med. 1997;45:803–9.

27. Layte R, Whelan CT. Explaining Social Class Inequalities in Smoking: The Role of Education, Self-Efficacy, and Deprivation. Eur Sociol Rev. 2009;25:399–410.

28. Covvey JR, Johnson BF, Elliott V, Malcolm W, Mullen AB. An association between socioeconomic deprivation and primary care antibiotic prescribing in Scotland. J Antimicrob Chemother. 2014;69:835– 41.

29. Murphy M, Byrne S, Bradley CP. Influence of patient payment on antibiotic prescribing in Irish general practice: a cohort study. Br J Gen Pract. 2011;61:e549–55.

30. Item -ICGP Web Site.

31. Viktil KK, Blix HS, Eek AK, Davies MN, Moger TA, Reikvam A. How are drug regimen changes during hospitalisation handled after discharge: a cohort study. BMJ Open. 2012;2:e001461.

32. Corsonello A, Pedone C, Corica F, Incalzi RA. Polypharmacy in elderly patients at discharge from the acute care hospital. Ther Clin Risk Manag. 2007;3:197.

33. Hummers-Pradier E, Himmel W, Kochen MM, Sorns U, Hummers-Pradier E. Drug changes at the interface between primary and secondary care. Artic Int J Clin Pharmacol Ther. 2004. https://doi.org/10.5414/CPP42103.

34. Karapinar F, Van Den Bemt PMLA, Zoer J, Nijpels G, Borgsteede SD. Informational needs of general practitioners regarding discharge medication: Content, timing and pharmacotherapeutic advices. Pharm World Sci. 2010;32:172–8.

35. Coll S, Walsh ME, Fahey T, Moriarty F. Hospital initiation of benzodiazepines and Z-drugs in older adults and discontinuation in primary care. Res Soc Adm Pharm. 2022;18:2670–4.

36. Pérez T, Moriarty F, Wallace E, McDowell R, Redmond P, Fahey T. Prevalence of potentially inappropriate prescribing in older people in primary care and its association with hospital admission: Longitudinal study. BMJ. 2018;363:k4524.

37. Lam KD, Miao Y, Steinman MA. Cumulative Changes in the Use of Long-Term Medications: A Measure of PrescribingComplexity. JAMA Intern Med. 2013;173:1546–7.

38. Falster MO, Charrier R, Pearson SA, Buckley NA, Daniels B. Long-term trajectories of medicine use among older adults experiencing polypharmacy in Australia. Br J Clin Pharmacol. 2021;87:1264–74.

